# Prone positioning of patients with moderate hypoxia due to COVID-19: A multicenter pragmatic randomized trial [COVID-PRONE]

**DOI:** 10.1101/2021.11.05.21264590

**Authors:** M Fralick, M Colacci, L Munshi, K Venus, L Fidler, H Hussein, K Britto, R Fowler, B Da Costa, I Dhalla, R Dunbar-Yaffe, L Branfield Day, T MacMillan, J Zipursky, T Carpenter, T Tang, A Cooke, R Hensel, M Bregger, A Gordon, E Worndl, S Go, K Mandelzweig, L Castelluci, D Tamming, F Razak, AA Verma, COVID Prone Study Investigators

## Abstract

**Objectives:** To assess the effectiveness of prone positioning to reduce the risk of death or respiratory failure in non-critically ill patients hospitalized with COVID-19

**Design:** Pragmatic randomized clinical trial of prone positioning of patients hospitalized with COVID-19 across 15 hospitals in Canada and the United States from May 2020 until May 2021.

**Settings:** Patients were eligible is they had a laboratory-confirmed or a clinically highly suspected diagnosis of COVID-19, required supplemental oxygen (up to 50% fraction of inspired oxygen [FiO2]), and were able to independently prone with verbal instruction. (NCT04383613).

**Main Outcome Measures:** The primary outcome was a composite of in-hospital death, mechanical ventilation, or worsening respiratory failure defined as requiring at least 60% FiO2 for at least 24 hours. Secondary outcomes included the change in the ratio of oxygen saturation to FiO2 (S/F ratio).

**Results:** A total of 248 patients were included. The trial was stopped early on the basis of futility for the pre-specified primary outcome. The median time from hospital admission until randomization was 1 day, the median age of patients was 56 years (interquartile range [IQR] 45,65), 36% were female, and 90% of patients were receiving oxygen via nasal prongs at the time of randomization. The median time spent prone in the first 72 hours was 6 hours total (IQR 1.5,12.8) for the prone arm compared to 0 hours (0,2) in the control arm. The risk of the primary outcome was similar between the prone group (18 [14.3%] events) and the standard care group (17 [13.9%] events), odds ratio 0.92 (95% CI 0.44 to 1.92). The change in the S/F ratio after 72 hours was similar for patients randomized to prone compared to standard of care.

**Conclusion:** Among hypoxic but not critically patients with COVID-19 in hospital, a multifaceted intervention to increase prone positioning did not improve outcomes. Adherence to prone positioning was poor, despite multiple efforts. Subsequent trials of prone positioning should aim to develop strategies to improve adherence to awake prone positioning.

**What is already known on this topic:** Prone positioning is considered standard of care for mechanically ventilated patients who have severe acute respiratory distress syndrome. Recent data suggest prone positioning is beneficial for patients with COVID-19 who are requiring high flow oxygen. It is unknown of prone positioning is beneficial for patients not on high flow oxygen.

**What this study adds:** Prone positioning is generally not well tolerated and innovative approaches are needed to improve adherence. Clinical and physiologic outcomes were not improved with prone positioning among hypoxic but not critically ill patients hospitalized with COVID-19.

## INTRODUCTION

As of July 2021, over 4 million people worldwide have died from Coronavirus disease 2019 (COVID-19). The strongest risk factors for death are older age, comorbid disease, and severity of presenting illness, most commonly the presence of hypoxia (1–3). Patients who present to hospital with severe hypoxia were typically cared for in an intensive care unit (ICU) with mechanical ventilation. Patients without severe hypoxia were commonly cared for on a hospital ward with supplemental oxygen via nasal prongs or face mask. However, approximately 20% of such patients progress to respiratory failure requiring mechanical ventilation(1,3).

In February of 2020, reports emerged that prone positioning of COVID-19 patients with severe hypoxia may reduce the risk of respiratory failure and death(4,5). Prone positioning has been part of clinical practice since the 1970s and is considered standard of care for mechanically ventilated patients who have severe acute respiratory distress syndrome (6,7). There are multiple physiologic reasons why prone positioning can improve oxygenation including decreased forces on the lungs from the heart and gastrointestinal organs that allow for improved lung expansion and decreased ventilation and perfusion mismatch (6). Early uncontrolled studies suggested prone positioning might also be beneficial for patients with COVID-19 who were not yet intubated, preventing the need for intubation(4,8). Because these findings occurred at a time when some intensive care units were overwhelmed and there were no effective treatments for COVID-19, they were shared widely on social media and in the lay press, leading to substantial adoption in practice.

Multiple observational studies have examined the effectiveness of prone positioning in non-intubated individuals with COVID-19(6). The results have been conflicting, with some studies showing a modest improvement in oxygenation and others showing no improvement (6). One randomized trial of 30 non-intubated patients with COVID-19 identified no improvement in oxygenation, however the trial was stopped early due to a lack of intervention adherence(9). A recently published meta-trial of patients requiring high flow nasal cannula identified a lower risk of treatment failure (i.e., a composite of intubation or death) for patients randomized to prone positioning(10). It remains unclear whether prone positioning is effective in managing patients with milder forms of hypoxia. Because prone positioning has potential risks to patients (e.g., aspiration, patient discomfort) and healthcare providers (i.e., requires more personnel time spent in the room with an infectious patient) and may be difficult for patients to tolerate, randomized trials are needed to evaluate the risks and benefits. We conducted a multicenter pragmatic randomized clinical trial to assess the effectiveness of prone positioning to reduce the risk of death or respiratory failure in non-critically ill patients hospitalized with COVID-19 (NCT04383613).

## METHODS

### TRIAL DESIGN

We conducted an unblinded pragmatic randomized clinical trial of prone positioning of patients hospitalized with confirmed or suspected COVID-19 in 15 hospitals in Canada and the United States from May 2020 until May 2021. Patients were eligible if they had a laboratory-confirmed or clinically highly suspected diagnosis of COVID-19, required supplemental oxygen (up to 50% FiO_2_), and were able to independently adopt a prone position with verbal instruction. Randomization occurred within 48 hours of hospitalization and patients were excluded if prone positioning was contraindicated (e.g., recent abdominal surgery), impractical (e.g., dementia, severe delirium), or mechanical intubation was indicated at the time of randomization as per the patient’s treating physician (Appendix). Co-enrollment in other clinical trials was allowed.

The trial protocol was approved by the institutional review board at each site (or by a centralized institutional review board as applicable) and was overseen by the trial’s steering committee (NCT04383613). An independent data monitoring committee was established and reviewed the interim analysis results so that the trial investigators were blinded. Informed consent was obtained from each patient. Patients were not involved in the design or planning of this study.

### DATA COLLECTION

We collected the following baseline data for each patient: demographics, comorbid conditions, vital signs, laboratory values, and imaging reports. We also recorded oxygenation and FiO_2_ data up to five times per day for the first 72 hours after randomization. Given the pragmatic nature of this trial, this data was abstracted from the patient chart based on routine vital signs documentation.

Self-reported time spent in the prone position was assessed from the time of randomization to 72 hours, and from 72 hours until day 7. The duration of the hospitalization, occurrence of adverse events, receipt of mechanical ventilation and vital status at hospital discharge were also recorded.

### RANDOMIZATION AND TRIAL PROCEDURES

Patients were randomly assigned in a 1:1 ratio, stratified by site to either prone positioning or standard of care (i.e., no instruction to prone position) using a web-based system with concealment of allocation. Patients were followed until the first of death, hospital discharge or 30 days. Patients randomized to prone positioning were recommended to adopt a prone position four times per day (up to 2 hours for each session) and encouraged to sleep in prone position overnight. These practices were recommended for up to 7 days in hospital, until hospital discharge, or until the patient no longer required supplemental oxygen (whichever came first). Patients were called by research coordinators at least twice during their time in hospital to encourage adherence to the study arm to which they were randomized to. Following a review of preliminary adherence data in December 2020, research coordinators were asked to remind patients daily of the arm they were randomized to. In the intervention arm, the patient’s attending physician and nurse provided similar reminders to patients and an order for prone positioning was entered into the electronic medical record at sites where possible.

### OUTCOME MEASURES

The primary outcome was a composite of in-hospital death, mechanical ventilation (i.e., intubation or bilevel positive airway pressure), or worsening respiratory failure defined as requiring at least 60% FiO2 for more than 24 hours. Given the inclusion of patients who had a pre-specified do-not-intubate order, the latter was included in our composite outcome. Secondary outcomes included: the components of the composite analyzed individually; time spent in prone position; change in the ratio of oxygen saturation (SpO_2_) to FiO_2_ (S/F ratio); time to recovery (defined as being on room air for at least 24 hours); time-to-discharge from hospital and the rate of serious adverse events.

### STATISTICAL ANALYSIS

Our trial was planned prior to effective treatments being available for patients with COVID-19. We planned for 80% power and a two-sided alpha of 0.05, and assumed the risk of our composite outcome would be 45%(1,11). With the goal of detecting a 15% reduction in the primary outcome, our total estimated sample size was 340 and allowed for 1 interim analysis at 50% patient recruitment (N=170).

The primary analysis was based on an intention-to-treat approach. Our primary outcome controlled for age and sex in a multivariable logistic regression model. Relative risk was estimated from the odds ratio. We planned *a priori* the following subgroup analyses of the primary outcome: (1) severity of hypoxemia at randomization based on arterial blood gas, (2) age, (3) chest radiograph findings and (4) amount of supplemental oxygen at baseline prior to randomization (Appendix).

Our secondary analysis of time-to-hospital discharge was analyzed using a Cox proportional hazards model that adjusted for age and sex. The change in the S/F ratio over the first 72 hours was analyzed using an ANCOVA model that adjusted for baseline S/F ratio as well as age and sex. We also conducted a *post-hoc* analysis to identify if longer time spent prone was associated with improved outcomes (Appendix).

The results from the interim analysis were reviewed by our data monitoring committee on May 4, 2021 while the trial investigators were blinded to the results. Upon reviewing results from the first 170 patients, the independent data monitoring committee requested to review results for all available patients after enrollment reached 230 patients. On May 10, 2021, they recommended stopping the clinical trial due to futility. Patients who were in the study at that time continued in the arm to which they were randomized.

## RESULTS

Of the 570 patients who were assessed for eligibility, 257 were randomized and 248 had a primary outcome recorded and were included in the intention-to-treat analysis (Figure 1). The median time from hospital admission until randomization was 1 day, 98% had a laboratory PCR confirmed diagnosis of COVID-19, the median age of patients was 56 years (IQR 45,65), 36% were female, 40% had hypertension, 27% had diabetes, and 11% had a diagnosis of chronic obstructive pulmonary disease (COPD) or asthma. Patients randomized to prone positioning were slightly older and more likely to have a diagnosis of hypertension while patients randomized to the control arm were more likely to be a current smoker or have a diagnosis of asthma or COPD at baseline (Table 1). The most common oxygen delivery method was nasal prongs (90%), the median oxygen saturation was 94% (IQR 93,96), and the median FiO_2_ was 32% (IQR 28,36). At baseline prior to randomization, 95% of patients received dexamethasone, 42% received remdesivir, and 1% received tocilizumab.

**Table 1.**
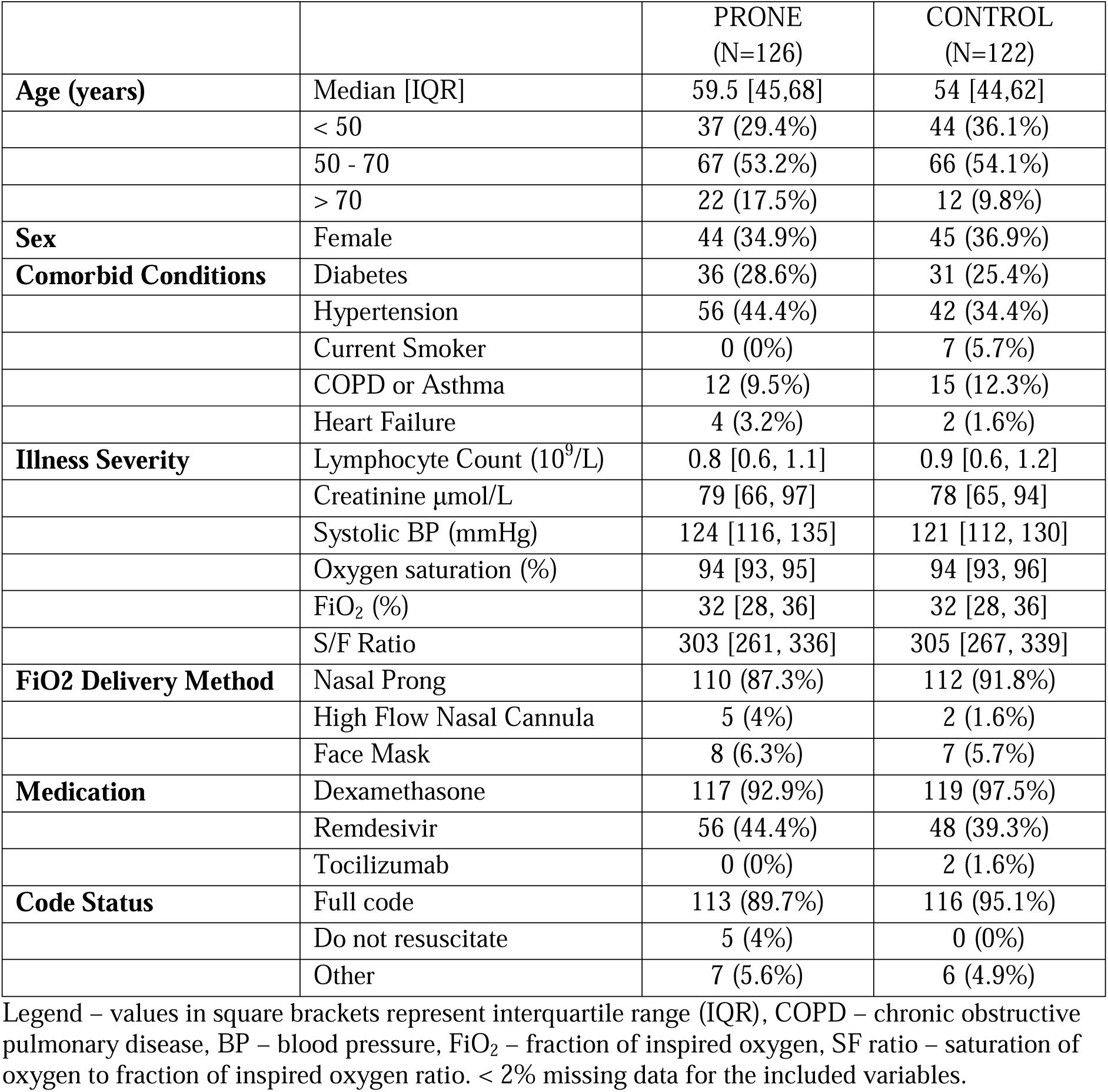
Baseline characteristics of included patients

|  |  | PRONE<br>(N=126) | CONTROL<br>(N=122) |
| --- | --- | --- | --- |
| **Age (years)** | Median [IQR] | 59.5 [45,68] | 54 [44,62] |
|  | < 50 | 37 (29.4%) | 44 (36.1%) |
|  | 50 - 70 | 67 (53.2%) | 66 (54.1%) |
|  | > 70 | 22 (17.5%) | 12 (9.8%) |
| **Sex** | Female | 44 (34.9%) | 45 (36.9%) |
| **Comorbid Conditions** | Diabetes | 36 (28.6%) | 31 (25.4%) |
|  | Hypertension | 56 (44.4%) | 42 (34.4%) |
|  | Current Smoker | 0 (0%) | 7 (5.7%) |
|  | COPD or Asthma | 12 (9.5%) | 15 (12.3%) |
|  | Heart Failure | 4 (3.2%) | 2 (1.6%) |
| **Illness Severity** | Lymphocyte Count ($10^9/L$) | 0.8 [0.6, 1.1] | 0.9 [0.6, 1.2] |
| | Creatinine $\mu\text{mol/L}$ | 79 [66, 97] | 78 [65, 94] |
|  | Systolic BP (mmHg) | 124 [116, 135] | 121 [112, 130] |
|  | Oxygen saturation (%) | 94 [93, 95] | 94 [93, 96] |
|  | FiO<sub>2</sub> (%) | 32 [28, 36] | 32 [28, 36] |
|  | S/F Ratio | 303 [261, 336] | 305 [267, 339] |
| **FiO2 Delivery Method** | Nasal Prong | 110 (87.3%) | 112 (91.8%) |
|  | High Flow Nasal Cannula | 5 (4%) | 2 (1.6%) |
|  | Face Mask | 8 (6.3%) | 7 (5.7%) |
| **Medication** | Dexamethasone | 117 (92.9%) | 119 (97.5%) |
|  | Remdesivir | 56 (44.4%) | 48 (39.3%) |
|  | Tocilizumab | 0 (0%) | 2 (1.6%) |
| **Code Status** | Full code | 113 (89.7%) | 116 (95.1%) |
|  | Do not resuscitate | 5 (4%) | 0 (0%) |
|  | Other | 7 (5.6%) | 6 (4.9%) |

Legend – values in square brackets represent interquartile range (IQR), COPD – chronic obstructive pulmonary disease, BP – blood pressure, FiO<sub>2</sub> – fraction of inspired oxygen, SF ratio – saturation of oxygen to fraction of inspired oxygen ratio. < 2% missing data for the included variables.

**Figure 1.**
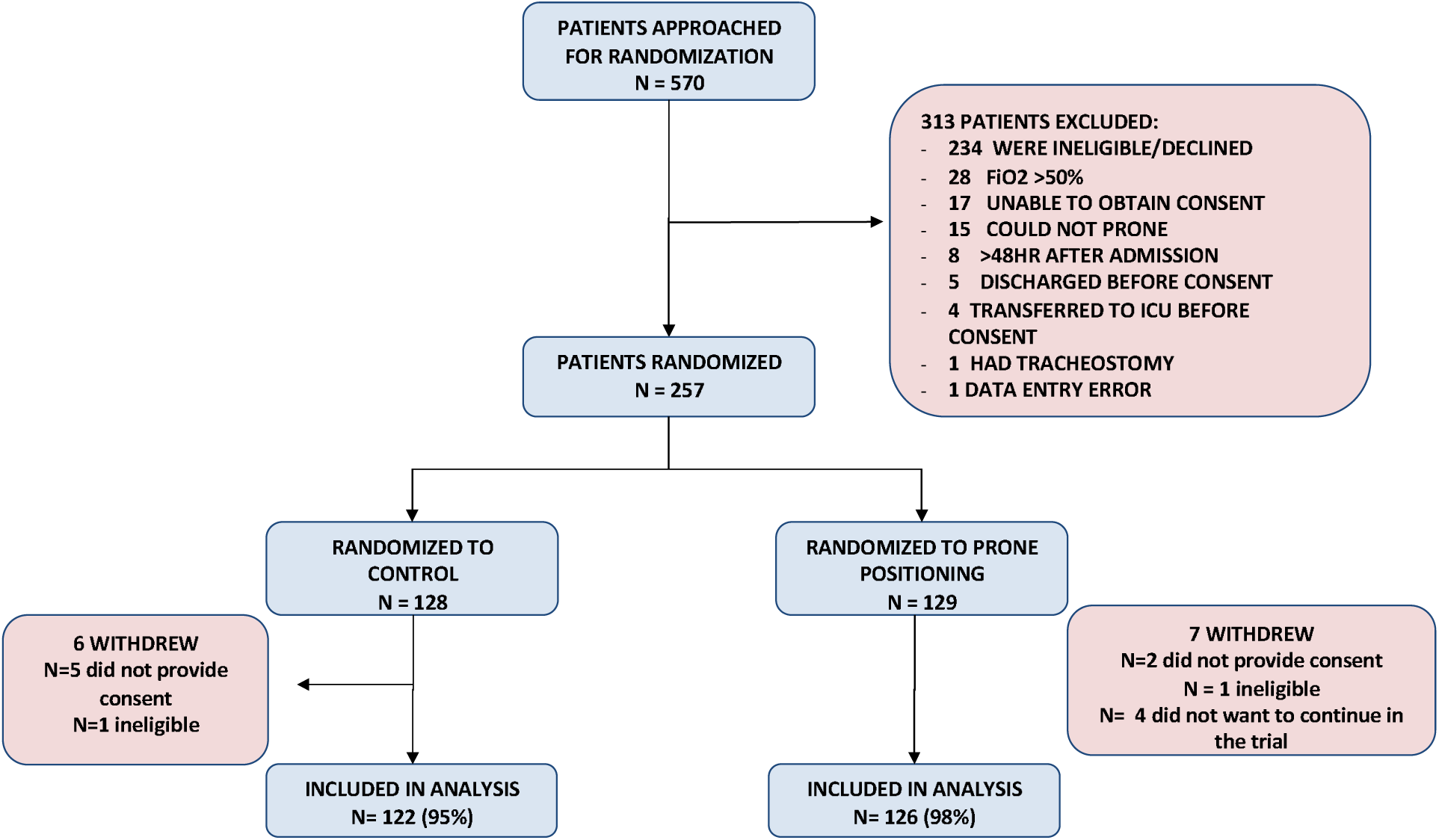
Enrollment and Randomization. FiO_2_ – fraction of inspired oxygen. Among the patients who withdrew due to lack of consent, this occurred because our trial allowed for deferred consent. Among the patients who withdrew because they were ineligible, that was because one patient was on room air at the time of randomization and the other patient was on > 50% FiO_2_ at the time of randomization.

Of the patients randomized to prone positioning, the median total time spent in prone position up to the first 72 hours was 6 hours [IQR 1.5,12.8] and 0 hours [IQR 0,2] in the control arm. After accounting for hospital discharge within the first 72 hours, on a per day basis this equated to approximately 2.5 hours per day in the prone arm compared to 0 hours per day in the control arm in the first 72 hours.

The median time from randomization until the primary outcome was 1 day. The rate of the primary outcome was similar between the prone group (18 [14.3%] events) and the standard of care group (17 [13.9%] events), odds ratio 0.92 (95% CI 0.44 to 1.92). In the subgroup analysis stratified by baseline hypoxia, the odds ratio for the primary outcome for patients randomized to prone compared to standard of care was 1.77 (95% CI 0.69,4.80) for those on more than 30% FiO_2_ and 0.26 (95% CI 0.05,0.96) for those on up to 30% FiO_2_. In the subgroup analysis that stratified by age, the odds ratio for the primary outcome for patients randomized to prone compared to standard of care were: 1.52 (95% CI 0.43,5.55) for those 55 years and younger and 0.74 (95% CI 0.30, 1.81) for those older than 55 years. In our *post-hoc* analysis stratified by adherence, for patients at hospitals with higher adherence, the odds ratio of the primary outcome was 0.98 (95% CI 0.26,3.73) whereas it was 0.96 (95% CI 0.38, 2.37) for patients at hospitals with lower adherence. The time spent prone at the high adherence sites in the prone group was 4 hours per day compared to 1 hour per day at the sites with lower adherence.

The adjusted difference in the S/F ratio between the time of randomization until 72 hours was similar between the two groups (Figure 2). The median time-to-hospital discharge was 5 days (IQR 3,9) for the prone arm and 4 days (IQR 3,8) for the control arm. Serious adverse events were rare and affected 5 (4.0%) of patients in the prone group and 3 (2.5%) patients in the standard of care group (Table 2).

**Table 2.** Primary and secondary outcomes

| **Primary outcomes\*** | **Prone** | **Control** | **Difference** |
| --- | --- | --- | --- |
| Composite of death, mechanical ventilation, and FiO2 greater than 60 | 18 (14.3%) | 17 (13.9%) | OR: 0.92 (0.44, 1.92) |
| Components of the composites |  |  |  |
| Death | 1 (0.8%) | 1 (0.8%) |  |
| Mechanical ventilation | 6 (4.8%) | 5 (4.1%) |  |
| FiO2 > 60% | 18 (14.3%) | 17 (13.9%) |  |
| **Secondary outcomes** | **Prone** | **Control** | **Difference** |
| S/F ratio after 72 hours** | 336 [216, 438] | 336 [232, 443] | MD: 8 (-18, 35) |
| Change in S/F ratio in first 72 hours**<br>(Median [IQR]) | 14 [-52,94] | 49 [-32,102] | MD: 20 (-17, 36) |
| Days to discharge (Median [IQR]) | 5 [3,9] | 4 [3, 8] | HR: 0.91 (0.69, 1.2) |
| Discharged | 115 (91.3%) | 118 (96.7%) |  |
| **Serious adverse events** | **Prone** | **Control** |  |
| Serious adverse event composite | 5 (4%) | 3 (2.5%) |  |
| Components of serious adverse events |  |  |  |
| Aspiration pneumonia | 2 (1.6%) | 1 (0.8%) |  |
| Venous thromboembolism | 3 (2.4%) | 2 (1.6%) |  |
| Other | 0 (0%) | 0 (0%) |  |
| **Hours spent prone** | **Prone** | **Control** |  |
| First 72 hours (Median [IQR]) | 6 [1.5, 12.8] | 0 [0, 2] |  |
| Missing first 72 hours (%) | 8 (6.3%) | 5 (4.1%) |  |
| From 72 hours to 7 days (Median [IQR]) | 0 [0, 12] | 0 [0, 0] |  |
| Missing 72 hours to up to 7 days (%) | 13 (10.3%) | 12 (9.8%) |  |

Legend: \* adjusted for age and sex, \*\* adjusted for baseline S/F ratio as well as age and sex. IQR – interquartile range, S/F ratio – saturation of oxygen to fraction of inspired oxygen ratio. OR – odds ratio. MD – mean difference. HR – hazard ratio.

**Figure 2.**
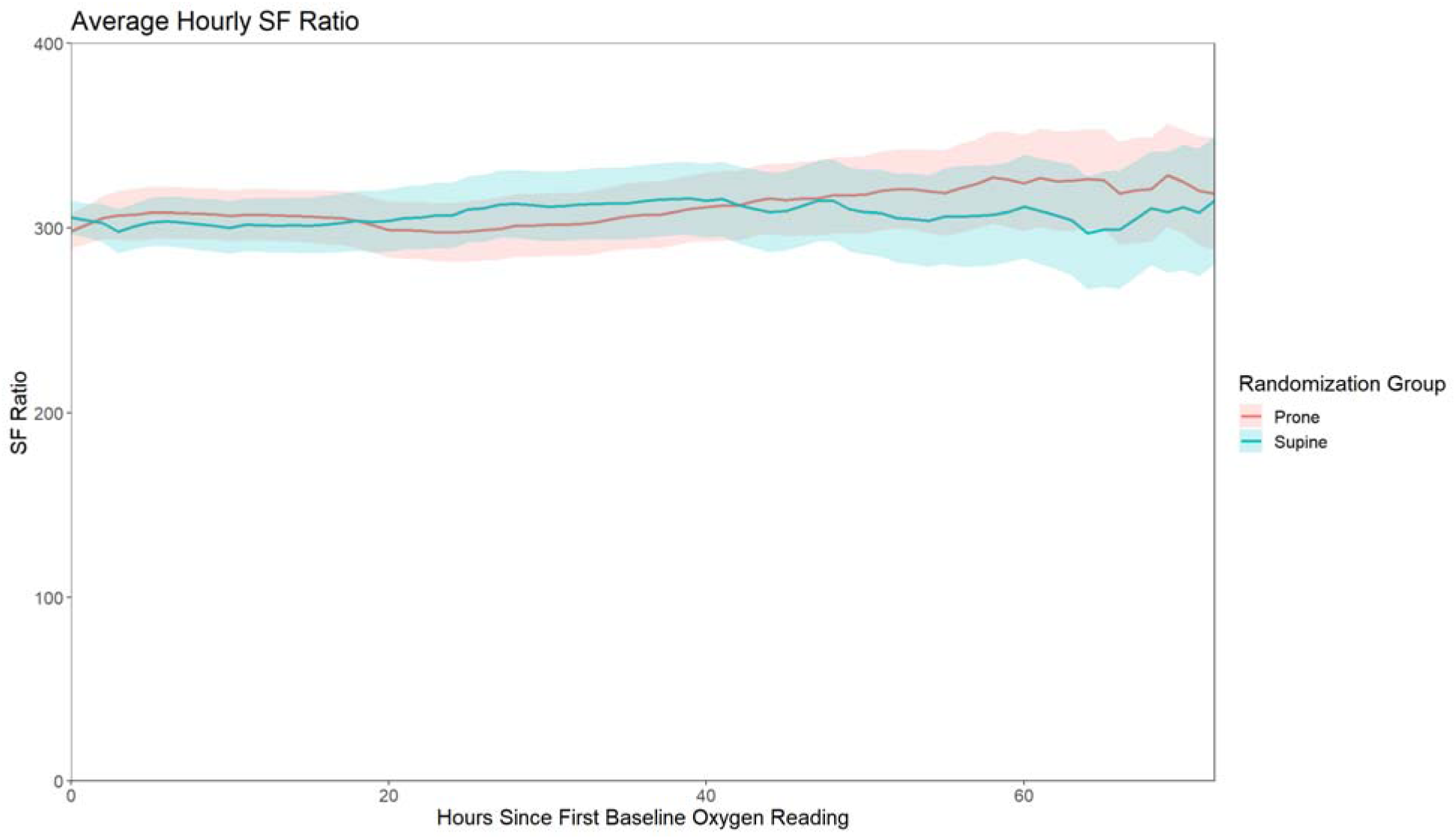
Change in SF ratio over time.

## Discussion

In this multicenter pragmatic randomized clinical trial of encouraging prone positioning in patients hospitalized with COVID-19 who were hypoxic but not critically ill, we did not observe improvements in the risk of the composite of death, mechanical ventilation, or worsening respiratory failure. Further, we did not observe improvements in oxygenation. However, the overall time spent prone per day was lower than planned despite inclusion and exclusion criteria identifying patients most likely to be able to prone and interventions to encourage adherence.

Initial reports suggested prone positioning might be potentially life-saving for patients hospitalized with COVID-19(12). However, early data were based on anecdotal reports and small case series. In the largest available studies of patients on a medical ward the duration of prone positioning was often not reported (6), and when reported, was typically less than 3 hours per day(6). Most of these studies focused on oxygenation rather than clinical outcomes, and identified improvements in oxygenation while the patient was prone but improvements did not necessarily persist after prone positioning was stopped(9). The one available randomized trial for patients with mild hypoxia showed no improvement in oxygenation, but the trial was small and stopped early because of poor adherence (i.e., mean duration of 1.6 hours spent prone in the first 72 hours)(13). Our study found that patients did not adhere to prolonged time spent prone, and there was no sustained improvement in oxygenation related to prone positioning. Specifically, the change in S/F ratio was similar between the two groups over the first 72 hours. Further, the rate of worsening respiratory failure, intubation, or death was similar between the two groups.

The time spent prone in our study can be characterized as “low intensity prone positioning”. We did not intend this *a priori*, and instead our study planned for four 2-hour prone sessions each day and prone sleeping at night. However, despite inclusion and exclusion criteria aimed at identifying people able to prone independently and with reminders to patients by nurses, physicians, and study team members, this was not achieved. The most common reason for the lack of adherence was patient discomfort. This feedback, coupled with the results from our study, confirm that simply instructing patients to lie prone and providing them with reminders is insufficient for most patients to spend a prolonged period in the prone position.

The low adherence to prone positioning in our study is important to contrast with prone positioning of patients who are in the ICU. Time spent prone in the ICU can be directly controlled by the healthcare team because the patients who are receiving invasive mechanical ventilation are typically sedated and potentially also receiving neuro-muscular blockade.

Benefits of prone positioning in ICU patients with ARDS were not observed in earlier trials that had shorter durations of prone positioning (i.e., less than 12 hours per day) (14). In a meta-trial (N=6 individual trials) of prone positioning patients with COVID-19 on high flow nasal cannula (median FiO2 at randomization of 60%) the median duration of prone positioning per day was less than five hours per day with the exception of one site in Mexico where it was 8.6 hours per day(10). Only the trial in Mexico identified a lower relative risk of the primary outcome, and the overall meta-analysis of the primary outcome identified a lower risk of the primary outcome for patients randomized to prone position(10). Additional trials of prone positioning of ward patients will help to identify if there are alternative strategies to adhere to prone positioning in the non-intubated patient and whether there is a sustained physiologic benefit across a certain subgroup that translates to improved clinical outcomes (e.g., non-intubated patients with a higher severity of acute respiratory failure). Based on the adherence we observed in our trial, innovative, more directive strategies may be needed to encourage awake patients to adopt a prone position for more than a few hours each day.

To understand how longer duration of time spent prone is associated with our primary outcome, we conducted a *post-hoc* exploratory analysis and compared outcomes at the sites with the highest adherence to prone positioning compared to the lowest adherence sites. We found no difference in the primary outcome at the sites with the highest adherence, but it is important to note that because of the relatively low number of overall events our null finding might be related to lack of statistical power or that the longer duration prone (approximately 3 more hours per day) was still insufficient for a clinically important benefit. We also conducted a pre-planned subgroup analysis to identify how baseline hypoxia may affect the efficacy of prone positioning. In that analysis we identified a lower risk of the primary outcome for patients randomized to prone positioning requiring 30% FiO_2_ or lower at the time of randomization. However, this should be considered hypothesis-generating because it was not pre-specified and because the low overall number of events which increases the possibility of chance alone underlying our observed findings. Future studies are needed to identify whether this finding is replicable and robust across other protocols.

Our study has several strengths. First, our study was pragmatic, multicenter, and included both academic and community hospitals. We anticipate our results reflect the effectiveness of real-world interventions to encourage prone positioning in similar healthcare settings. Second, we collected a physiologic outcome (i.e., oxygenation), clinically relevant outcomes (i.e., death or mechanical ventilation), included patients with a do-not-resuscitate order to enhance generalizability, and surrogates for healthcare utilization (e.g., need for more than 60% FiO_2_ and length of hospital stay). Third, unlike most of the available published studies, our trial included data on the potential risks of prone positioning.

The most important limitation of our study was poor adherence to time spent prone. As described, this likely reflects the real-world challenges of lying in a prone position when sick with a respiratory and multisystem viral illness, and without high nurse-to-patient ratios. Future studies are needed to determine whether a greater amount of time spent in the prone position is associated with clinical benefit including whether specific devices or interventions (e.g., smartphone apps, pillows/padding, prone positioning teams) can increase time spent prone. Second, our expected event rate was lower than anticipated because our study was planned prior to effective treatments being available. These treatments (e.g., dexamethasone, remdesivir, tocilizumab), at least in part, resulted in a lower event rate that we anticipated and thus larger randomized trials would be needed to identify a smaller absolute risk reduction. Third, we lacked data on PPE use and the number of times a healthcare provider had to enter the room to help reposition a patient to aid with prone positioning. Both are important to evaluate when considering the potential drawbacks of prone positioning. Fourth, FiO2 for non-intubated patients depends on multiple physiologic factors and needs to be estimated for certain devices (e.g., a non-rebreather mask) based on the flow rate of oxygen delivered.

## CONCLUSION

In our multicenter international pragmatic trial, we did not observe improved clinical outcomes, or physiologic outcomes with prone positioning among hypoxic but not critically ill patients hospitalized with COVID-19. The trial was stopped early based on the futility of finding the pre-specified effect size. Ongoing studies are evaluating whether prone positioning might be beneficial for non-intubated patients with more severe forms of hypoxia. The poor adherence to prone positioning we observed highlights that it is generally not well tolerated and innovative approaches are needed to improve adherence.

## Supporting information

COVID Prone Study Investigators (Group Authors)

## Data Availability

Please reach out to the authors with any requests to access data produced in the present study.

## Appendix Additional details for analysis

We had planned to adjust for age, sex, and hospital. Because of fewer than anticipated events (i.e., patients who experienced the primary outcome) our model only adjusted for age and sex. We also planned to report odds ratios, risk ratios, and risk differences. Because our trial was null we reported odds ratios alone. We were unable to perform the planned subgroup analysis based on severity of hypoxemia at randomization because few patients had an arterial blood gas performed, or the analysis based on chest radiograph findings because nearly all patients had an abnormal chest x-ray.

### Post-hoc analysis

To avoid immortal time bias that could occur using an approach that conditions on time spent prone after randomization, we calculated the rate of the primary outcome among patients randomized to prone compared to control at the hospitals with the highest adherence to prone positioning compared to those with the lowest adherence.

### COI

Dr. Jonathan Zipursky has received payments for medicolegal opinions regarding the safety and effectiveness of drugs outside the submitted work. Dr. Amol Verma is Provincial Clinical Lead for Quality Improvement in General Internal Medicine at Ontario Health and an AMS Healthcare Fellow in Compassion and Artificial Intelligence. He has received funding for COVID-related research from CIHR, Canadian Frailty Network, St. Michael’s Hospital, Sinai Health System, and St. Michael’s Hospital Foundation. Dr. Mike Fralick is a consultant for a start-up company that has developed a point of care diagnostic test for COVID-19 using CRISPR. Dr. Fahad Razak has received an award from the Mak Pak Chiu and Mak-Soo Lai Hing Chair in General Internal Medicine, University of Toronto, outside the submitted work and is an employee of Ontario Health.

### Ethics

The study was approved by Clinical Trials Ontario (CTO) ethics committee (reference number 3184), as well as the respective ethics boards of participating centres not governed by CTO (listed below).

1. Beth Israel Deaconess Medical Center (2020P000648)
2. Northwestern Memorial Hospital (STU00212937-CR0001)
3. Sault Area Hospital (2020-12-01)
4. Scarborough Health Network (COV-20-009)

## Acknowledgements

We thank Dr. Art Slutsky and Dr. Kevin Thorpe who were our data monitoring committee. We also thank the research ethics boards and contracts departments who provided expedited review of our trial so that we can launch expeditiously in the setting of the COVID-19 pandemic. Finally, and most importantly, we are deeply grateful to the trial participants chose to participate in a clinical trial while enduring this serious illness.

